# Relapse timing is associated with distinct evolutionary dynamics in DLBCL

**DOI:** 10.1101/2023.03.06.23286584

**Authors:** Laura K. Hilton, Henry S. Ngu, Brett Collinge, Kostiantyn Dreval, Susana Ben-Neriah, Christopher K. Rushton, Jasper C.H. Wong, Manuela Cruz, Andrew Roth, Merrill Boyle, Barbara Meissner, Graham W. Slack, Pedro Farinha, Jeffrey W. Craig, Alina S. Gerrie, Ciara L. Freeman, Diego Villa, Michael Crump, Lois Shepherd, Annette E. Hay, John Kuruvilla, Kerry J. Savage, Robert Kridel, Aly Karsan, Marco A. Marra, Laurie H. Sehn, Christian Steidl, Ryan D. Morin, David W. Scott

## Abstract

Diffuse large B-cell lymphoma (DLBCL) is cured in over 60% of patients, but outcomes are poor for patients with relapsed or refractory disease (rrDLBCL). Here, we performed whole genome/exome sequencing (WGS/WES) on tumors from 73 serially-biopsied patients with rrDLBCL. Based on the observation that outcomes to salvage therapy/autologous stem cell transplantation are related to time-to-relapse, we stratified patients into groups according to relapse timing to explore the relationship to genetic divergence and sensitivity to salvage immunochemotherapy. The degree of mutational divergence increased with time between biopsies, yet tumor pairs were mostly concordant for cell-of-origin, oncogene rearrangement status and genetics-based subgroup. In patients with highly divergent tumors, several genes acquired exclusive mutations independently in each tumor, which, along with concordance of genetics-based subgroups, suggests that the earliest mutations in a shared precursor cell constrain tumor evolution. These results suggest that late relapses commonly represent genetically distinct and chemotherapy-naïve disease.

## Introduction

DLBCL is an aggressive and heterogeneous lymphoma for which standard-of-care R-CHOP (rituximab with cyclophosphamide, vincristine, doxorubicin, and prednisone) immunochemotherapy results in long term remission in more than 60% of patients.^1^ However, outcomes are poor for the 30-40% of patients with primary refractory or relapsed disease (rrDLBCL) even after salvage therapy and autologous stem cell transplant (ASCT).^2,3^ The landscape of coding and non-coding somatic variants in DLBCL at diagnosis is well established,^4–8^ and several studies have examined the mutational landscape of cohorts of rrDLBCL to compare the post-treatment genetic landscape to that of diagnostic DLBCL, identifying somatic variants that occur more frequently in rrDLBCL such as *MS4A1*, *TP53*, *NFKBIE*, *FOXO1*, *CREBBP*, and *KMT2D*.^9–12^ Although several of these mutations are prognostic at diagnosis for the likelihood of relapse, they are insufficient to explain the poor outcomes experienced by rrDLBCL patients.

Tumor evolution is usually considered to follow one of two models: linear or branching evolution. Linear evolution is defined when the relapse tumor alone harbors a set of exclusive variants not found at diagnosis, implying direct descent of the relapse from the diagnostic tumor. Branching evolution is characterized by exclusive variants in both diagnostic and relapse tumors. In the transformation of follicular lymphoma (FL) to aggressive DLBCL (tFL), this branching pattern of evolution is considered evidence of a persistent common precursor cell (CPC) that is ancestral to both lymphomas.^13,14^ To-date, studies of DLBCL tumor evolution have leveraged circulating tumor DNA (ctDNA) and/or limited targeted capture space to examine the evolutionary dynamics of relapse in small cohorts, providing some evidence that branching evolution predominates even when tumor pairs are genetically very similar.^9,15–17^ However, the degree to which persistent CPC populations might contribute to DLBCL relapse is not yet known.

Critically, no studies have yet examined the evolution of the mutation landscape together with overall tumor biology, which can be evaluated through gene expression profiling (GEP)-based cell-of-origin (COO)^18,19^ and dark-zone signature (DZsig)^20,21^ classification. More recently, genetics-based classifiers have been developed that leverage co-occurrence of somatic variants to identify shared biology within DLBCL. Intriguingly, the three studies that described genetics-based groups converged on 5-7 highly overlapping subgroups.^7,8,22–24^ The LymphGen algorithm is currently the only publicly-available tool for assigning genetics-based subgroups to an individual biopsy.^22^ These classification systems are becoming the foundation for precision medicine in DLBCL, and while the current assumption is that the features that underlie the classification of each tumor would be fixed over time, this requires formal testing.

Here, we examined a large population-based cohort of rrDLBCL and confirmed that response rate and outcomes to salvage (immuno)chemotherapy and ASCT are superior for patients with late relapses relative to primary refractory or early relapse. To examine the genetic and evolutionary relationships between diagnostic and rrDLBCL underlying these clinical differences, we assembled a cohort of 129 patients with multiple DLBCL biopsies, and interrogated them with a combination of fluorescence *in situ* hybridization (FISH) for recurrent rearrangements, GEP for COO and DZsig, and/or whole genome (WGS, 68 patients) or whole exome sequencing (WES, 5 patients) of two or more tumors per patient. Clonal evolution analyses showed an association between the time to relapse and genetic divergence, with late relapses exhibiting a pattern of deep branching evolution. This also revealed an unexpected pattern of convergent evolution among divergent tumor pairs. Our findings of divergent evolution in late relapses suggests these are effectively *de novo* aggressive disease, therefore retaining chemosensitivity and driving superior outcomes in this group.

## Results

### Late relapses have superior outcomes

Considering prior observations that outcomes to salvage therapies are related to progression/relapse timing,^25,26^ we first sought to confirm this observation in a large population-based patient cohort (“outcomes cohort”). We identified 221 patients with *de novo* DLBCL treated with front line R-CHOP(-like) therapy that experienced DLBCL progression or relapse (Table S1-2). All patients received salvage chemotherapy (89% received GDP (gemcitabine, dexamethasone, cisplatin) +/- rituximab)^27^ with intention-to-treat with consolidative ASCT in patients with (immuno)chemotherapy responsive disease. Patients were categorized into three relapse timing categories: primary refractory disease was defined as progression or relapse within 9 months of diagnosis, approximating 3 months post-end of treatment and consistent with the definition provided by Hitz et al.^28^ Late relapses were defined as more than 24 months after diagnosis, with this timing reflecting the definition of EFS24 – a validated end point in which patients event-free 24 months following immunochemotherapy collectively have superior disease-related outcomes.^29^ Early relapses were defined as those occurring between 9-24 months from diagnosis. We found significant differences in both response rates (Figure 1A) and the proportion of patients who ultimately received consolidative ASCT (Figure 1B), demonstrating superior (immuno)chemosensitivity of tumors of patients experiencing late relapses. Patients experiencing late relapse had significantly superior progression-free survival (PFS) and overall survival (OS) relative to patients that experienced primary refractory or early relapse when considering either time from first progression/relapse (Figure 1C-D) or from receipt of ASCT (Figure 1E-F). Outcome differences persisted after adjusting for age at diagnosis and IPI at relapse (Extended Data Figure 1). Differences in outcomes were driven by both the proportion of patients receiving ASCT and the outcomes following ASCT. While there was no significant difference between the proportion of patients with early and late relapse receiving ASCT, post-ASCT outcomes were significantly superior in the late relapse group. The overall superior outcomes from the time of progression in the early relapse vs primary refractory group was related to the greater propotion of patients receiving ASCT as there were no differences in post-ASCT outcomes between these groups.

**Figure 1.**
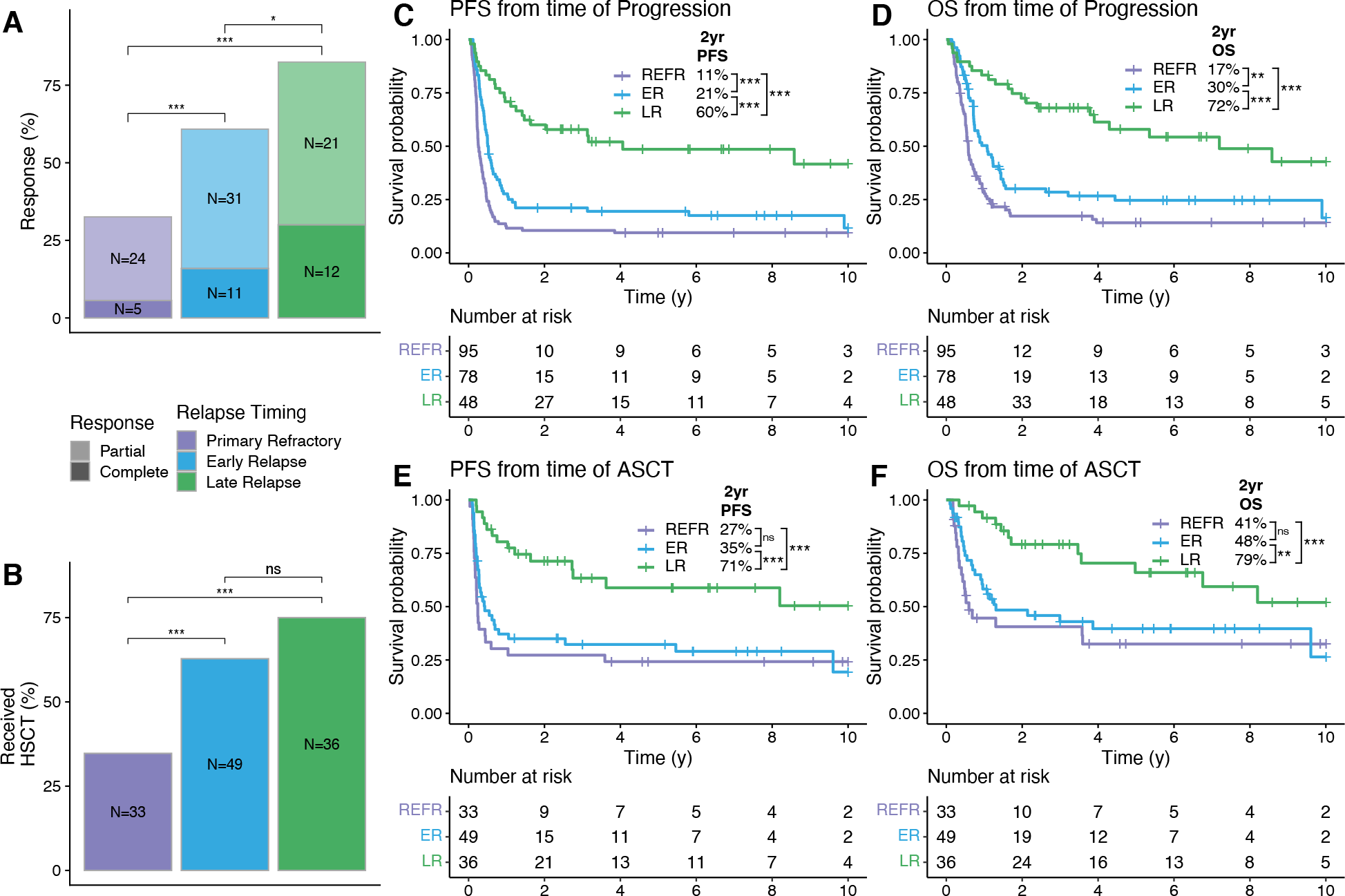
Relationship between relapse timing and outcomes to salvage therapy. **A, B.** The percent of patients in each relapse timing category whose relapse responded to salvage therapy (**A**) and who received ASCT (**B**). Groups were compared with pairwise Fisher’s exact tests. **C-F.** Kaplan-Meier curves showing PFS and OS from the time of progression or ASCT. P-values were determined with a log-rank test. * P < 0.05; ** P < 0.01; *** P < 0.001.

### Late relapses are highly divergent

To examine the underlying tumor biology and patterns of evolution driving the superior outcomes observed in late relapses, we assembled a cohort of patients that experienced relapse with available serial DLBCL biopsies (“molecular characterization cohort”). While a uniform curative treatment approach at time of relapse was important to demonstrate a relationship between relapse timing and (immuno)chemosensitivity in the outcomes cohort, the criteria for inclusion in the molecular characterization cohort was sufficient material from multiple biopsies (and matched constitutional DNA for WGS/WES). Thus, patients who were not treated with intention to consolidate with ASCT were included along with patients with prior indolent lymphoma as long as multiple DLBCL biopsies were available. The distribution of time to relapse in the molecular characterization cohort differs from that observed in the outcomes cohort, reflecting historic patterns of obtaining a biopsy to confirm relapse less frequently in primary refractory disease and availability of constitutional DNA. In total, 129 patients were identified, of which 29 had prior follicular lymphoma (FL; 22.5%), two had prior marginal zone lymphoma (MZL; 1.6%), and one had prior chronic lymphocytic leukemia (CLL; 0.8%). Among the 97 patients with apparently *de novo* DLBCL at diagnosis, 11 had a subsequent FL diagnosis (11.3%) and four had a subsequent MZL (including extranodal MZL of mucosa-associated lymphoid tissues (MALT); 4.1%). Six total patients had discordant low-grade bone marrow lymphoid infiltrates at diagnosis and two at relapse. Depending on tissue availability, each pair of biopsies was interrogated with a combination of FISH for *MYC*, *BCL2*, and/or *BCL6* rearrangements, digital GEP (NanoString DLBCL90) for COO and DZsig classification,^19,30^ and/or WGS or WES (Extended Data Figure 2 and Table S3-4). Of the 129 total patients, 73 had sufficient material for WGS (N=68) or WES (N=5) (Figure 2), and 21 were also included in the outcomes cohort (4 primary refractory, 7 early relapses, and 10 late relapses). Of these 73 patients, two late relapse patients had prior DLBCL biopsies that were not available for sequencing, while the remainder of patients had the first and at least one subsequent DLBCL biopsy sequenced.

**Figure 2.**
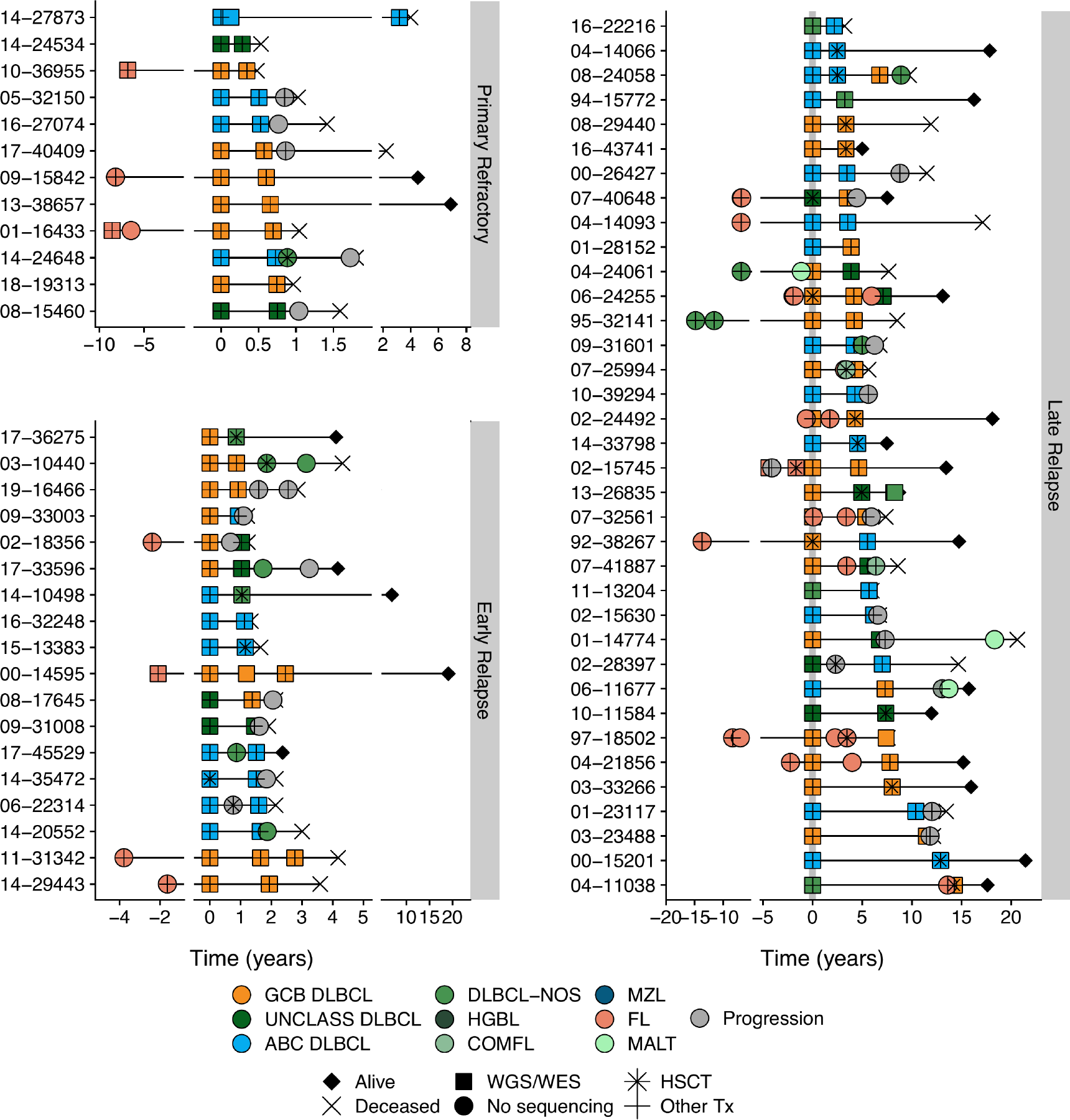
Sequencing cohort patient histories. Disease and treatment histories for all known biopsies and progressions for patients for which WGS/WES data were generated, distributed according to relapse timing categories. Five patients were omitted due to incomplete histories. DLBCL tumors are colored according to NanoString COO where available or are otherwise labeled DLBCL-NOS (not otherwise specified). COMFL, composite lymphoma with areas of DLBCL and FL morphology; FL, follicular lymphoma; MZL, marginal zone lymphoma; MALT, extranodal MZL mucosa-associated lymphoid tissue; HGBL, high-grade B-cell lymphoma; PROG, clinical progression without a biopsy.

The use of FFPE tissues for most samples resulted in variable sequencing depth (mean 48.6X across WGS samples and 97X in exomes; Extended Data Figure 3A and Table S5). Although tumors in the late relapse category had significantly lower depth of coverage on average, there was no correlation of total mutation burden with coverage (Extended Data Figure 3B) indicating that sequencing depth was sufficient to comprehensively detect clonal variants. We also performed deep targeted DNA sequencing of a panel of genes relevant for LymphGen classification (“LySeqST”, Table S6) on multiple biopsies subjected to WGS from 47 patients and on a single biopsy from another 15 patients. The LySeqST assay achieved a mean depth of 812X across its capture space (Extended Data Figure 4A and Table S7). The lower variant allele frequencies (VAFs) of variants detected by LySeqST alone vs. genomes demonstrates that it enhanced detection of subclonal variants that fall below the limit of detection of WGS (Extended Data Figure 4B).

Next, we explored the overall divergence of mutations by comparing the number of shared (common between both biopsies) and exclusive (present in only one biopsy) mutations between the first two DLBCL biopsies in each patient. For this and all subsequent analyses, we pooled the LySeqST and WGS variant calls and only retained variants at positions with a sequencing depth of at least 10 unique molecules in all tumors from the same patient. While primary refractory and early relapse tumors have a rich landscape of variants shared between tumors (Figure 3A middle), many late relapses have few, with most mutations exclusive to either the diagnostic or relapse biopsy (Figure 3A top and bottom). In both primary refractory and early relapse disease, the number of mutations shared between tumors is strongly correlated with the total number of variants identified at either diagnosis or relapse with slopes nearing unity, demonstrating that most variants are shared between tumors (Figure 3B). In contrast, the number of shared variants in late relapses is only weakly correlated to the total mutation burden in either tumor (Figure 3B). Comparing the percentage of exclusive variants in each tumor to the time between biopsies reveals a clear linear trend, where tumor pairs separated by many years have very few shared variants (Figure 3C and Table S8). This trend is consistent when considering time to relapse as a categorical variable (Extended Data Figure 5), when the absolute number of exclusive mutations is considered (Extended Data Figure 6), and is independent of genome coverage (Table S9). The linear relationship between exclusive variants and relapse timing was also consistent when transformed FL tumors were considered separately from *de novo* DLBCL (Extended Data Figure 7). These results are consistent with a branching evolution model of evolution, where late relapse tumor pairs arise from a distant common ancestor harboring few lymphoma-defining mutations.

**Figure 3.**
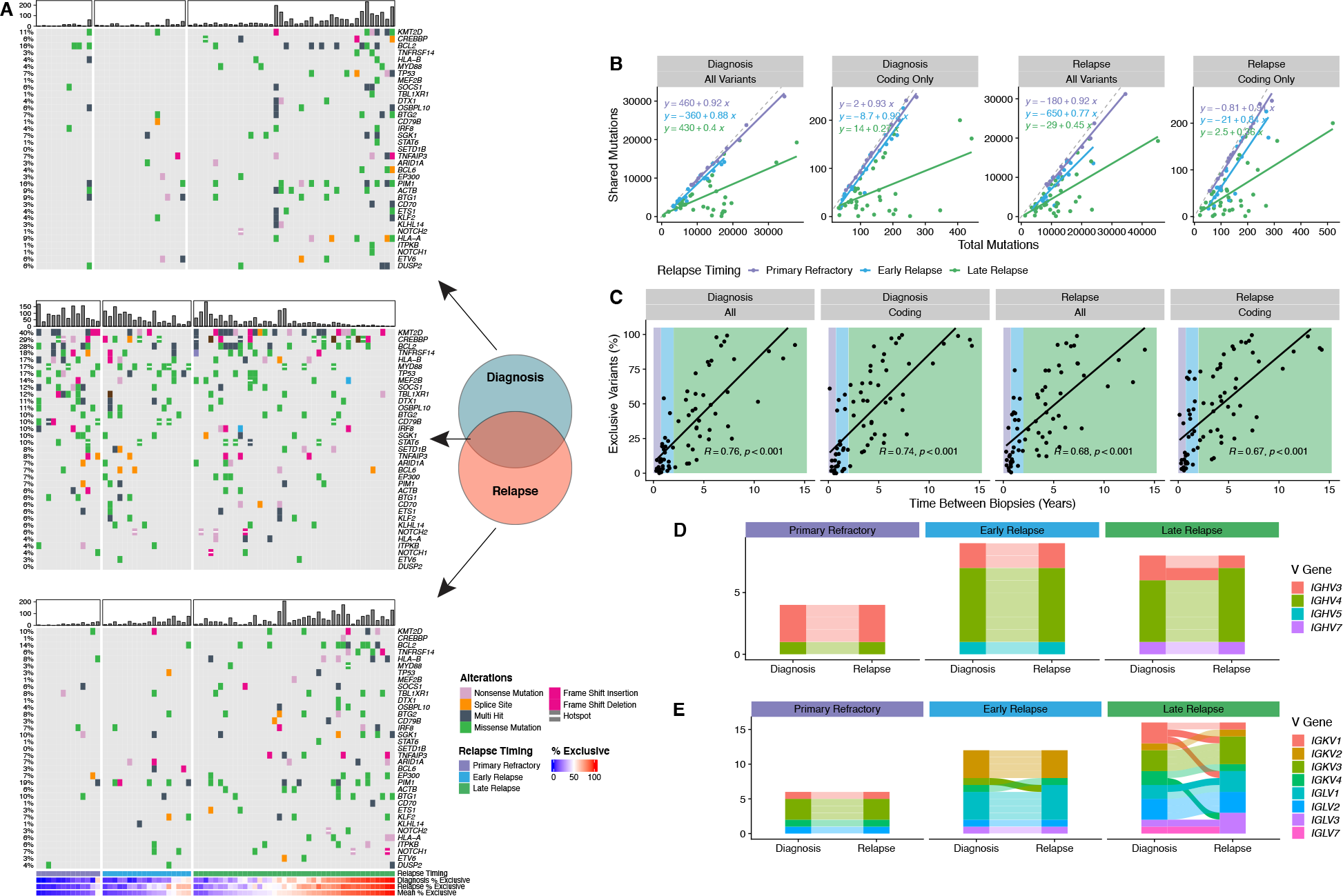
Patterns of evolution in diagnostic and relapse tumor pairs. **A** Oncoplots of variants identified exclusively at diagnosis (top), relapse (bottom), or shared between biopsies (middle), highlighting the most frequently mutated genes involved in LymphGen classification. Patients are stratified by relapse timing and ordered by the mean percentage of exclusive variants. Barplots indicate the number of coding mutations present per patient in each mutation subset. **B** The relationship between total variants (all or coding only) at diagnosis or relapse vs. the number of mutations shared between tumors. The dashed grey line represents the line of unity. **C** The percent of variants exclusive to either diagnostic or relapse tumors as a function of time between biopsies. R represents the Pearson correlation coefficient. **D, E.** Concordance of heavy chain (**D**) and light chain (**E**) V gene usage derived from RNAseq data for tumor pairs colored by V gene subgroup. In all plots, alluvia connecting each tumor pair are opaque for discordant pairs and translucent for concordant pairs. *N.B.* Where V gene usage was discordant but both V genes belong to the same subgroup, the color is consistent across timepoints.

Given the high degree of divergence observed in some late relapse tumors, we used RNAseq data to identify functional expressed IG receptor rearrangements and confirm clonal relatedness of tumor pairs (Table S8). All 4 primary refractory and 9 early relapse patients had concordant IGHV gene usage, while one of 8 late relapses was discordant (Figure 3D). Light chain rearrangements were more frequently discordant, which may suggest ongoing receptor editing (Figure 3E).^31^ The lone patient with discordant heavy chain rearrangements also had discordant light chain rearrangements, suggesting these tumors are not clonally related and the second DLBCL is effectively *de novo*.

### Temporal dynamics of structural variants

Rearrangements of *MYC*, *BCL2*, and *BCL6* are important drivers of aggressive lymphoma biology and contribute to disease and genetics-based classification.^22,32,33^ *BCL2* rearrangement status was concordant in all patients tested (Figure 4A and Table S3), consistent with the established origin of *BCL2* rearrangements during V(D)J recombination in early B cell differentiation.^34^ In 26 patients where WGS identified *BCL2* breakpoints in two or more tumors, the breakpoints were always identical (Table S11).

**Figure 4.**
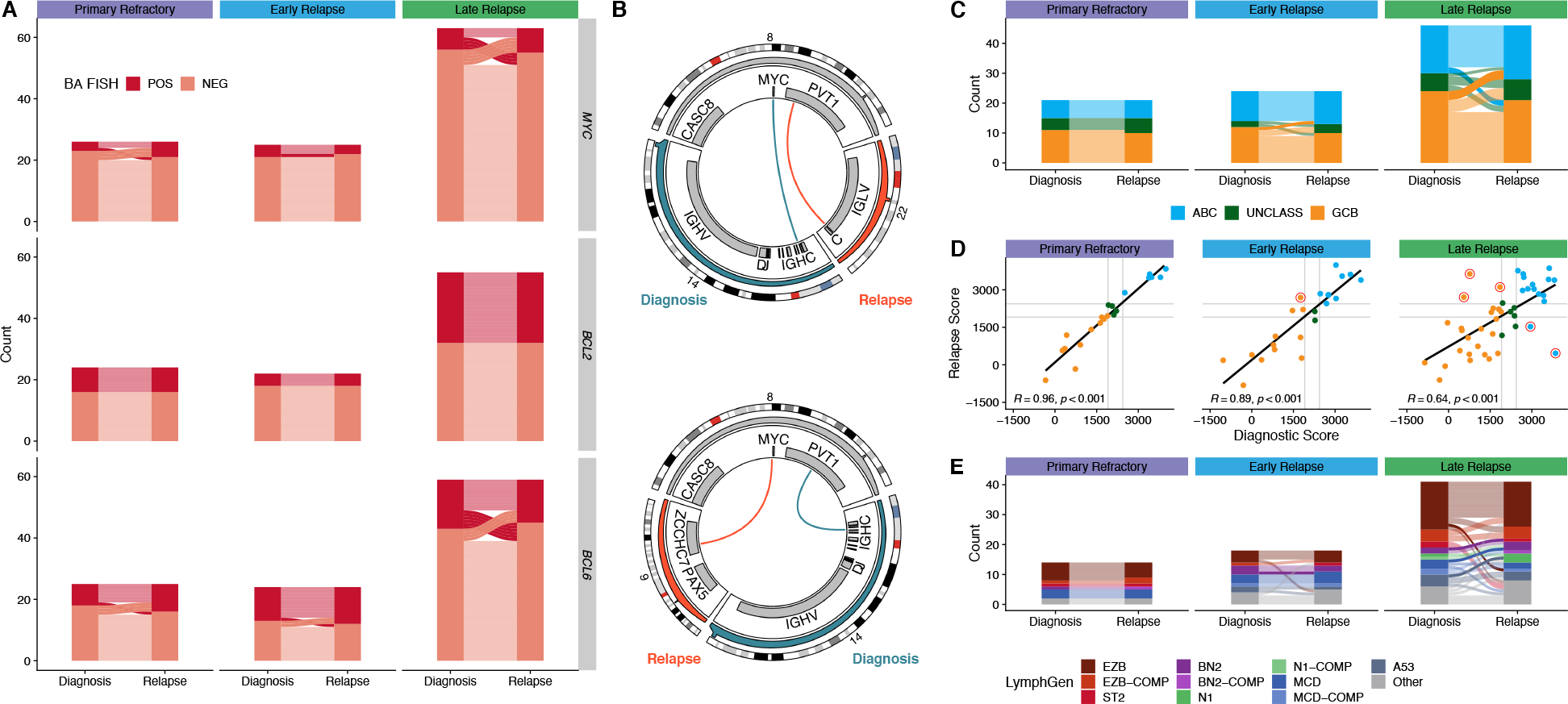
Comparison of structural variants and GEP and genetic classifications between biopsies. **A.** Concordance of BA-FISH results between diagnosis and first relapse for *MYC*, *BCL2*, and *BCL6* translocations. **B.** Circos plots showing discordant *MYC* translocations in two patients who experienced late relapse. Top: a tumor pair that was positive for BA-FISH at both timepoints; bottom: a tumor pair that was BA-FISH postive at diagnosis and negative at relapse. **C.** Alluvial comparison of COO classifications in diagnostic/relapse pairs stratified by relapse timing. Frank discordance (ABC to GCB or *vice versa*) is indicated by opaque alluvia. **D.** A scatter plot comparing DLBCL90 COO scores across tumor pairs. Red circles highlight frank discordance in COO classification. R values indicate Pearson correlation coefficient. **E.** Comparison of LymphGen classifications between tumor pairs. Frank discordance (a switch between two mutually exclusive non-Other classifications) is emphasized with opaque alluvia.

In contrast, *MYC* rearrangement status was discordant between timepoints in 14/114 patients tested (12%), and for *BCL6* in 17/108 patients tested (16%) with BA FISH (Figure 4A). As a proportion of patients in which any tumor harbored a rearrangement, the rate of discordance is substantial, with 70% of 20 *MYC*-translocated patients discordant between tumors and 65% of 26 *BCL6*-translocated patients. Interestingly, in all 10 patients where *BCL6* rearrangements were identified by WGS at multiple timepoints, the breakpoints were identical. Persistent *BCL6*::IGH rearrangements were only found in primary refractory and early relapse, while all late relapses involved persistent *BCL6* rearrangements with non-IG partners.

*MYC* breakpoints were identified by WGS in multiple tumors from 6 patients, one of which was cryptic to FISH.^35^ Two patients, both late relapses, had different translocation partners at diagnosis and relapse (Figure 3B and Table S8), demonstrating independent acquisition of *MYC* translocations in each tumor on the background of the existing *BCL2* translocation in both of these patients, making all of these high-grade B-cell lymphoma with *MYC* and *BCL2* rearrangements. A third late relapse patient had an identical *BCL6*::*MYC* translocation in both tumors, and two patients with early relapse and one with primary refractory disease also had identical *MYC* breakpoints in both tumors. These findings suggest that in patients experiencing late relapse, the *MYC*-translocated aggressive lymphoma is effectively eradicated by treatment, while the indolent CPC harboring a *BCL2* translocation or other variants can persist for many years, and new *MYC* translocations may occur on development of a subsequent aggressive lymphoma. That these late relapses with discordant *MYC* rearrangements both harbored *MYC*::IGH translocations at diagnosis, while the late relapse with a concordant *MYC* rearrangement had a persistent *BCL6*::*MYC* translocation, is consistent with previous studies suggesting that *MYC* rearrangements involving the immunoglobulin (IG) loci have a more aggressive phenotype than those with non-IG rearrangements,^21,36^ indicating that a CPC may harbor such non-IG *MYC* rearrangements and acquire additional variants at relapse to reproduce the aggressive phenotype. Our findings indicate that *BCL6* rearrangements follow a similar pattern.

### Biological consistency of tumor pairs

We next evaluated the consistency of molecular subgroups using GEP and LymphGen over time. First, using digital GEP (the NanoString DLBCL90 assay)^19,20^ to compare COO and DZsig from 91 patients and considering only frank changes in COO classification (*i.e.* a switch from GCB to ABC or *vice versa*), we observed a high level of concordance between diagnosis and relapse (Table S3). None of 20 primary refractory patients, only one of 24 early relapse patients (4.2%), and only 5 of 47 late relapses (10.6%) were discordant (Figure 4C). Comparison of the NanoString linear predictor scores (LPS) between timepoints revealed a weaker correlation in late relapse patients, possibly indicating additional biological divergence not captured by this binary classification (Figure 4D). A similar trend was observed in DZsig scores applied to GCB or COO-unclassified tumors (Extended Data Figure 8).

To evaluate consistency in genetic subgroup assignment, we compared LymphGen classifications across diagnostic/relapse tumor pairs. In total, this yielded a genetic classification for 80% of tumors. In all relapse timing categories, LymphGen classifications were highly concordant, with discordance mainly occurring in patients with overlapping composite or “Other” (not assigned to any subgroup with sufficient confidence) classifications (Figure 4E, Table S12). However, there was a single early relapse patient out of 18 (5.6%) with a frank discordance (BN2 to MCD), and 4 discordant cases among 41 late relapses (9.8%). Thus, there was high consistency of both GEP and genetic classification despite the mutational divergence observed in late relapses.

### Convergent evolution in divergent pairs

The relative consistency of molecular subgroups as proxies for tumor biology is at odds with our observation that late relapses share relatively few mutations with the diagnostic tumor. To reconcile these differences, we performed phylogenetic analyses of all available tumors from each patient, leveraging all coding mutations alongside non-coding mutations in regions known to be affected by aberrant somatic hypermutation (aSHM; Table S13).^6^ In primary refractory tumors, the vast majority of somatic variants are found in the shared phylogenetic “trunk” (clonal in both tumors; Figure 5A). In early relapse tumors, the trunk is comparatively smaller and branching evolution gives rise to exclusive mutations in both diagnostic and relapse tumors (Figure 5B). In late relapses, few mutations are in the trunk, with substantial divergence on each branch (Figure 5C-D). In one patient described earlier, the trunk comprised a single shared coding mutation (Figure 5E). This patient had 7.5 years between diagnosis and relapse, is among only 5 tumors with frank discordance of LymphGen and COO classifications, and has different inferred IGH and IGK/L rearrangements (Figure 3D-E), together providing strong evidence that these tumors are not clonally related and thus arose independently.

**Figure 5.**
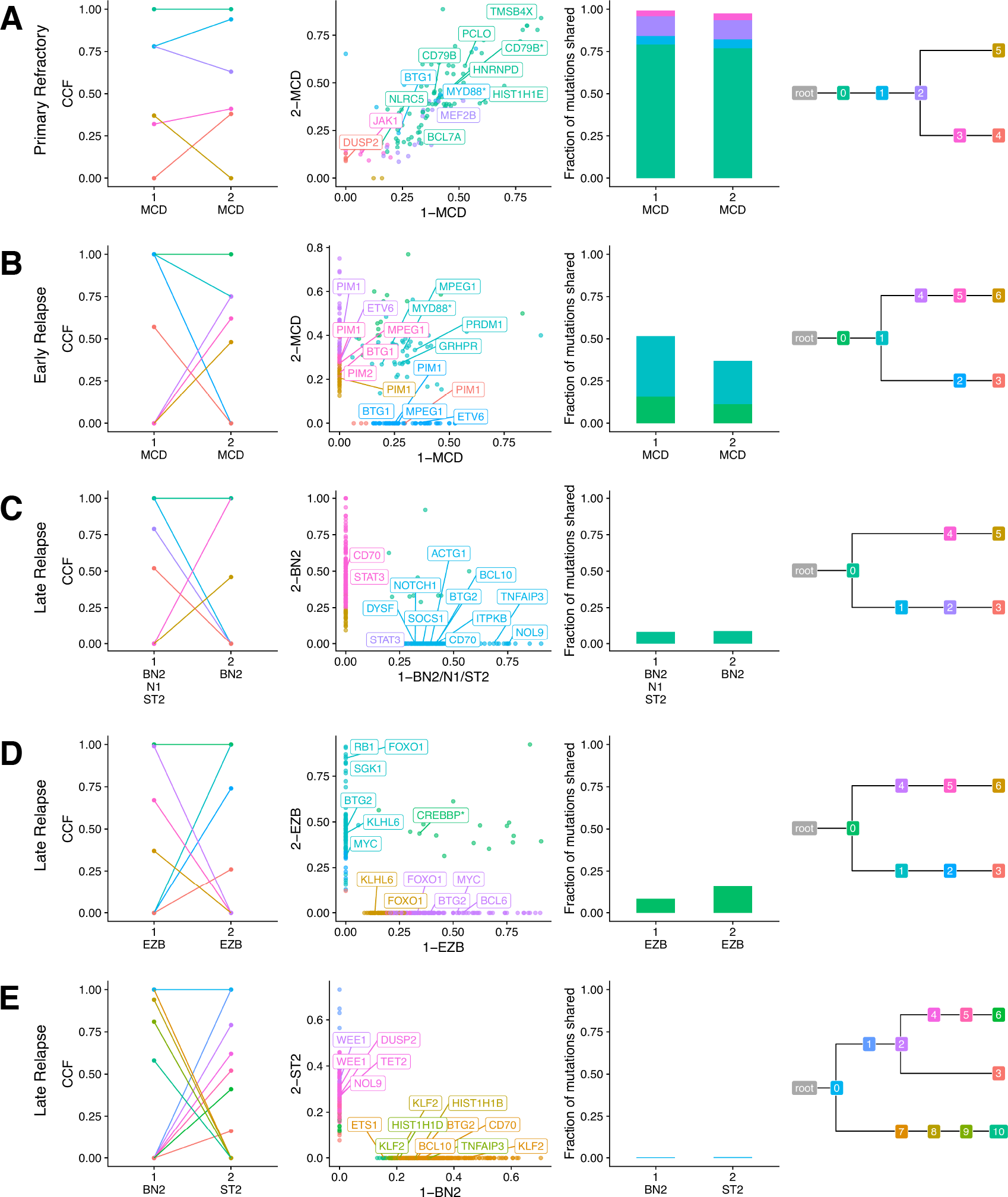
Representative phylogenetic reconstructions. Each row of plots displays data for a single patient. Tumors are labeled according to order of occurrence and LymphGen classification. Subclones are colored consistently across all plots for each patient. From left to right: Cancer cell fraction of subclones estimated by PhyClone; VAF of each variant as a scatter plot with the diagnostic tumor on the x-axis and relapse tumor on the y-axis with selected genes labeled; the fraction of mutations shared between both tumors (*i.e.* all mutations in a cluster with a CCF > 0.1); and the inferred phylogenetic relationship between tumors. Hotspot mutations at *MYD88* L265P and *CD79B* Y179 and missense mutations in the *CREBBP* lysine acetyltransferase (KAT) domain are indicated with an asterisk.

In addition, we noted a tendency for divergent tumor pairs to harbor exclusive mutations among the same genes. In the representative MCD-classified early relapse tumor pair, each tumor has independently acquired mutations in *BTG1*, *PIM1*, and *ETV6* (Figure 5B); the representative late relapse BN2 tumor pair in *CD70* and *STAT3* (Figure 5C); and the representative late relapse EZB tumor pair in *FOXO1*, *KLHL6*, *BTG2*, and *MYC* (Figure 5D). To examine this phenomenon more broadly, we used variant calls from 28 patients (2 early relapse, 26 late relapse) with divergent patterns of evolution, defined as having at least 25% of mutations exclusive to each tumor, and identified truncal (shared among all tumors from the same patient) and exclusive mutations. In total, 28 genes had two or more truncal mutations (Figure 6A and Table S14). Some genetic subgroup-defining mutations are among these, including *MYD88*^L265P^ mutations in 4/5 mutated patients and *CREBBP* KAT domain mutations in 5/5 mutated patients. Loci affected by aberrant somatic hypermutation (aSHM), including *BCL2*, *IGLL5*, and *BTG2* had a high number of both truncal and exclusive mutations, suggesting that aSHM can be an early shared event but continues after divergence. Other genetic subgroup-defining mutations were less frequently truncal, including *NOTCH2* PEST domain truncating mutations (1/3), *EZH2*^Y646^ (0/2), and *TET2* mutations (2/7). As individual mutations such as *EZH2*^Y646^ may be considered for treatment stratification or prognosis, this finding underscores the importance of re-characterizing late relapses.

**Figure 6.**
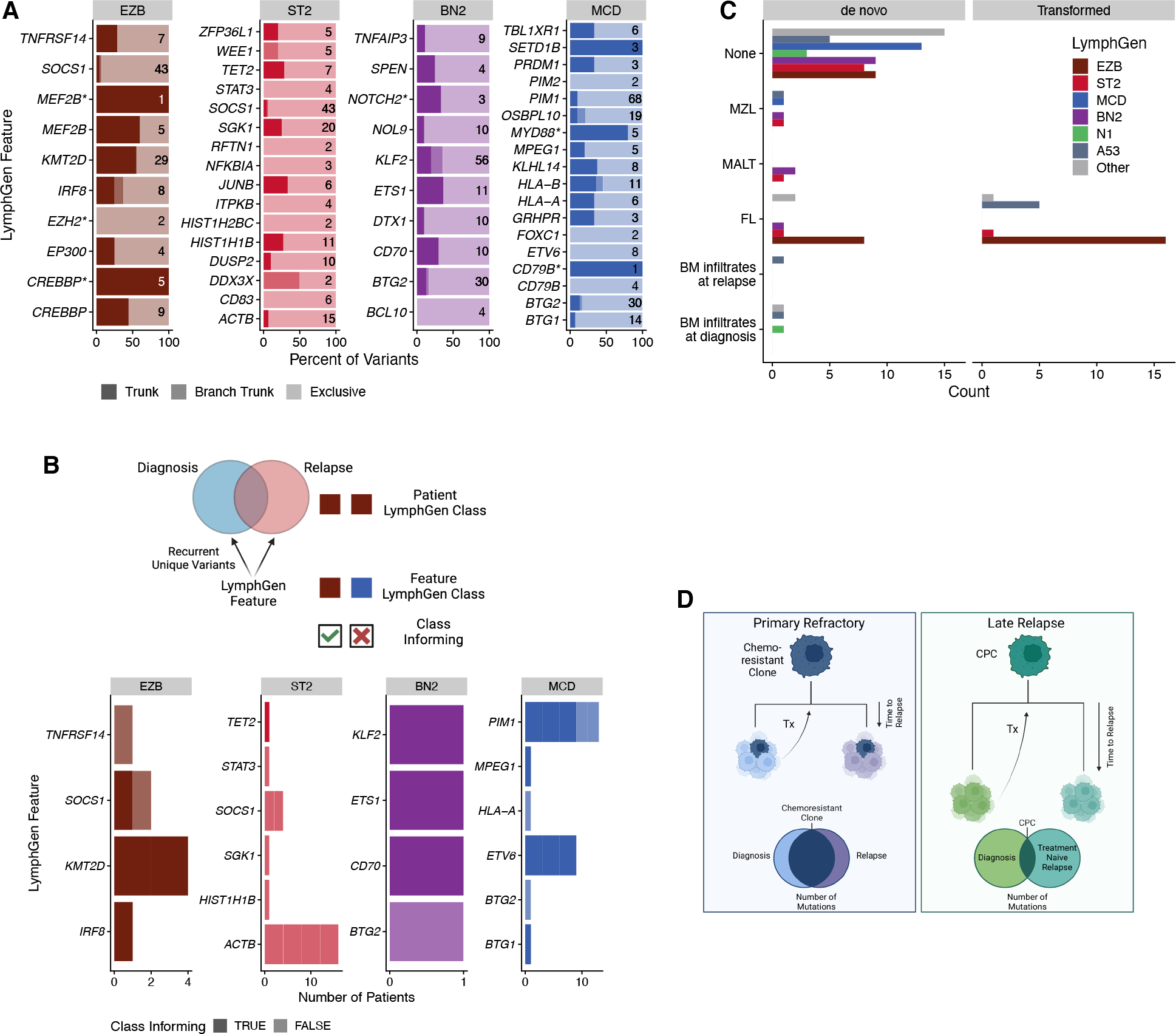
Classification features in divergent tumor pairs. **A.** LymphGen classification features that were mutated in two or more tumors from the same patient. Trunk variants (darkest) are identical in all tumors from the same patient; branch trunk are variants not shared across all tumors but are common between at least two; and exclusive are those found in only one tumor. Numbers on each bar represent the total number of variants considered in each feature. **B.** LymphGen classification features that acquired exclusive variants in two or more tumors from the same patient. Class informing indicates that the mutations arose in patients in which LymphGen classification matched the class association of the acquired variants. **C.** Patient LymphGen classifications stratified according to associated low-grade lymphoma entities. Transformed indicates low-grade disease preceded the first DLBCL diagnosis, while *de novo* indicates that the low-grade diagnosis was made after DLBCL diagnosis. **D.** A model of the relationship between relapse timing, evolutionary patterns, and outcomes. Created with BioRender.com

In total, 16 of 28 patients with divergent patterns of tumor evolution had tumors that independently acquired mutations in two or more of the same lymphoma-related genes (Table S15-16). *KMT2D*, *PIM1*, *ACTB*, and *ETV6* were among genes most frequently recurrently mutated in tumor pairs (Figure 6B). We compared LymphGen classifications of patients in which these mutations recurred to the LymphGen subgroup with which each feature is associated. Some features, such as MCD-related *PIM1* and *ETV6*, recurred in patients with MCD-classified tumors, while *ACTB*, associated with the ST2 LymphGen class, only recurred in patients without ST2-classified tumors (Figure 6B). Lastly, we examined the relationship between LymphGen classifications and prior or subsequent low-grade disease. The genetic similarities between different LymphGen classes and different low-grade mature B-cell lymphomas has been noted previously, and it has been speculated that this similarity reflects shared evolutionary history and CPC features in each LymphGen subgroup-indolent lymphoma pairing.^22,24^ As expected based on this model, patients with FL at any time in their disease course had DLBCL tumors predominantly classified as EZB, while the few MZL/MALT lymphomas occurred in patients with BN2- and ST2-classified DLBCLs (Figure 6C). Overall, our findings are consistent with a shared CPC origin for DLBCL at diagnosis, relapse and any prior or subsequent indolent disease, and indicate that the few mutations in the CPC population constrain the possible genetic features acquired in each tumor, resulting in biologically consistent disease even in patients where the DLBCLs share few somatic variants.

## Discussion

By leveraging multiple metrics of tumor evolution including cytogenetics, GEP, and unbiased genome- and exome-wide sequencing, we have established distinct patterns of tumor evolution that correlate strongly with the timing of DLBCL relapse. The high rate of mutations exclusive to both diagnostic and relapse biopsies shows that branching evolution predominates in late relapses, strongly supporting the existence of persistent CPC populations capable of giving rise to multiple DLBCL over time. However, GEP- and genetics-based classifications remained largely consistent, suggesting that the earliest clonal mutations in a CPC constrain the biology of the subsequent DLBCL(s). This constrained evolution may be the basis for the remarkable convergence of the three studies that have defined the genetic subgroups of DLBCL.^7,8,23,24^ Subgroup-defining mutations used in the LymphGen classification were sometimes among the inferred CPC mutations identified, while others were not consistently clonal, suggesting that additional genomic aberrations or other non-genetic features, such as DNA methylation or tumor microenvironment, that are responsible for establishing distinct CPC populations are still to be discovered. Furthermore, these CPC mutations appear to constrain the set of loci that acquire mutations during tumorigenesis. As some of these loci include regions affected by aSHM, constrained mutations are not always pathogenic drivers. However, aSHM is well known to target the transcriptional start sites of highly actively transcribed genes, including in normal memory B cells,^37,38^ and many aSHM loci have strong associations to genetic subgroups^22^, and therefore provide footprints of the phenotypic states cells have passed through *en route* to DLBCL.

Our observations of tumor evolution in DLBCL have both similarities to, and differences from, those made previously in studies of the transformation of indolent lymphomas, including CLL and FL. In Richter’s transformation (RT) of CLL, evolution follows a more linear pattern, and evidence for subclones that will eventually seed RT have been observed at diagnosis.^39^ Parry *et al.* compared RT to DLBCL using the Harvard genetics-based classification system and found that, while RTs clonally-unrelated to the preceding CLL clustered with *de novo* DLBCLs, the clonally-related RTs clustered separately, demonstrating a genetic uniformity to DLBCLs originating from a CLL-like CPC.^8,40^ However, the Harvard classification lacks a *NOTCH1*-driven subgroup, which is hypothesized to be most similar to CLL, so the relationship between RT and DLBCL genetic subgroups remains underexplored. In contrast, several studies of FL transformation have demonstrated branching patterns of evolution similar to what we have observed in DLBCL, where FL and tFL originate from a shared ancestral CPC, and no evidence of the eventual tFL-seeding subclone has been found in diagnostic FL samples.^13,14^ As expected based on genetic similarities and the proposed CPC origin of indolent and aggressive lymphomas, we demonstrated, for the first time, that DLBCLs were classified into the genetic subgroups with the most similarity to the indolent lymphoma diagnosed in each patient. Importantly, the entire spectrum of DLBCL genetic subgroups were observed in the late relapses of patients without any history of indolent disease. This demonstrates that distinct CPCs may provide the substrate for all genetic subgroups, and that patients do not have to manifest overt indolent disease in order for the DLBCL relapse to exhibit constrained evolution.

The patterns of DLBCL tumor evolution observed here help explain the responses to salvage (immuno)chemotherapy observed at disease relapse in DLBCL. In primary refractory disease, the pattern of tumor evolution suggests that innate chemoresistance is present at diagnosis, with little change in the composition of mutations upon treatment (Figure 6D). In this study and others, these tumors do not typically respond to further (immuno)chemotherapy-based salvage regimens and outcomes are poor,^3^ while alternatives to chemotherapy have been shown to produce superior outcomes in this patient population.^41,42^ The population of primary refractory patients should therefore be the focus in identifying both genetic and non-genetic mechanisms of resistance to front-line immunochemotherapy.

In contrast, our observations of the biology of late relapse are consistent with elimination of the original DLBCL but persistence of a CPC harboring a very small number of mutations. These CPC populations subsequently give rise to a genetically divergent DLBCL with a large number of newly-acquired mutations (Figure 6D). Although they share genetic features, the repertoire of driver mutations in the relapse is not preserved. As applications of precision medicine are being explored in DLBCL with emphasis on the rrDLBCL population, genomic analysis of relapsed samples is warranted. As these late relapses are effectively chemotherapy naïve, immunochemotherapy-based regimens may remain a rational treatment option.

## Data Availability

Complete sequencing data including WGS/WES, RNAseq, and targeted sequencing will be deposited at the European Genome/Phenome Archive (EGA) under study ID EGAS00001007053. All code for variant calling and phylogenetic analysis are available as part of LCR-modules (https://github.com/LCR-BCCRC/lcr-modules). An extensive repository of code for statistical analysis is available at https://github.com/LCR-BCCRC/DLBCL-tumor-evolution.

https://ega-archive.org/studies/EGAS00001007053

## Acknowledgements

This study was supported by a Large Scale Applied Research Project funded by Genome Canada (13124), Genome BC (271LYM), Canadian Institutes of Health Research (CIHR) (GP1-155873), the British Columbia Cancer Foundation (BCCF) and the Provincial Health Services Authority (PHSA). It was also supported by Terry Fox Research Institute (TFRI) Program Project Grants (1061, 1108), TFRI Marathon of Hope Cancer Centre Network, the Genome BC Marathon of Hope Cancer Centre program (MOH001), and grant 1P01CA229100 from the National Cancer Institute. RDM and CS are supported by Michael Smith Foundation for Health Research, Career Investigator Awards. DWS is supported by a Michael Smith Foundation for Health Research, Health Professional Investigator Award. BC is supported by a CIHR Canada Graduate Scholarship Doctoral Award.

## Online Methods

### Cohort selection

The BC Cancer lymphoma database was queried to identify all patients with DLBCL diagnosed between 2001-2020 with documented progression or relapse after front-line curative-intent R-CHOP-like chemoimmunotherapy. All patients were transplant-eligible and treated with salvage chemoimmunotherapy with intention to treat with autologous hematopoietic stem cell transplant. Patients with incidental discordant bone marrow involvement with low-grade B-cell lymphoma were included. Exclusion criteria were HIV positivity, transformed DLBCL, and isolated central nervous system (CNS) relapse. For most patients, the date of end of treatment was unavailable, so the definition of primary refractory as relapse or progression within 9 months of diagnosis is based on a typical 6 month course of treatment followed by progression or relapse within another 3 months^28^. This study was reviewed and approved by the University of British Columbia-BC Cancer Research Ethics Board in accordance with the Declaration of Helsinki.

For sequencing and other assays, BC Cancer lymphoma database was queried to identify patients with multiple biopsies of DLBCL morphology, extended to include patients with transformed lymphoma and more complex treatment histories. Six patients were identified from the Canadian Cancer Trials Group LY17 cohort. At Princess Margaret Cancer Centre, patients with relapsed DLBCL were prospectively accrued. Patients with a history of indolent disease were identified based on occurrence of indolent lymphoma (FL, CLL/SLL, MALT, MZL) or bone marrow infiltrates at any time in disease course. Samples were selected for sequencing on the basis of sufficient available material for two or more timepoints and peripheral blood for germline controls. Where insufficient material was available for sequencing, samples were subjected to fluorescence *in situ* hybridization (FISH) and/or gene expression profiling (GEP).

### Fluorescence *in situ* hybridization

Tissue Microarrays (TMAs) were constructed from duplicate 0.6 mm cores. FISH was performed on formalin-fixed paraffin-embedded (FFPE) material on whole tissue or TMA sections using commercially available Vysis LSI (Abbott) or Metasystems XL break apart probes for MYC (Abbott 01N63-020; Metasystems D-6030-100-TC), BCL2 (Abbott 05N51-020; Metasystems D-6018-100-OG) and BCL6 (Abbott 01N23-020; Metasystems D-6016-100-OG). Images were captured using a Metafer CoolCube 1 camera and Metafer software (version 3.11.8). Two independent observers scored 100 nuclei per biopsy, except for small or poor-quality biopsies where only 50-100 nuclei could be scored. Tumors in which <50 nuclei could be scored were considered a failed FISH result. For biopsies with discrepant scores, consensus was reached by a third observer. Rearrangement was defined as a break-apart signal in ≥5% of cells, and copy number variation was defined as ≥3 fused signals in ≥25% of cells.

### NanoString Gene Expression Profiling

RNA was extracted from FFPE biopsies using the Qiagen AllPrep DNA/RNA FFPE Kit (80234) or isolated from DNAse treatment of total nucleic acids (TNA) extracted using the ALINE FFPE GenePure Purification Kit (FP-2001-BC-384), automated on the Hamilton Nimbus robot. Digital gene expression profiling (GEP) was performed on RNA using the DLBCL90 assay on the nCounter platform (Nanostring), as previously described^19,20^.

### Library construction and sequencing

Samples were selected for sequencing on the basis of the availability of sufficient material for two or more biopsies along with peripheral blood for germline controls. The preservation of each tumor sample (FFPE or fresh frozen) is indicated in Table S1. For FFPE samples, total nucleic acid (TNA) was extracted using the ALINE FFPE GenePure Purification kit. When ≥ 300 ng of TNA was extracted, TNA was treated with S1 nuclease to reduce the rate of chimeric fragments derived from single-stranded DNA and overhangs^43^. DNA was isolated from TNA by RNase treatment. Paired-end DNA libraries for sequencing were generated using a PCR-based protocol as previously described^44^. For fresh frozen tissues, DNA was extracted and subjected to library construction as previously described^45^.

Targeted capture sequencing was performed using a panel of 126 genes designed specifically for LymphGen classification (“LySeqST”; Table S3). Hybridization capture was performed on the same library as WGS when possible, or else a new nucleic acid extraction from the same biopsy was obtained for library construction and capture.

### Alignment and variant calling

Genomes, exomes, and targeted capture data were aligned against the GRCh37 reference genome using bwa-mem 0.7.17^46^. To minimize the rate of FFPE artifacts, somatic variants were identified with the SLMS-3 pipeline, which employs an ensemble of Manta/Strelka2^47,48^, SAGE, MuTect2^49^, and LoFreq^50^, and variants called by three or more variant callers are passed. Somatic variants were annotated with VCF2MAF and the Ensembl Variant Effect Predictor^51^. Variant calls were “augmented”: all variant calls for a given patient were pooled, and read support for each variant was tallied up for each genome or exome from that patient, thereby recovering low-support variants that may be undetected by our stringent variant calling pipeline. When both WGS and LySeqST capture data were available, variant calls were pooled for each biopsy. A variant was excluded if there weren’t at least 10 unique reads covering its genomic position in all sequenced tumors from the same patient; no tumor had more than 30% dropout of variants using this method. LymphGen classification was performed with these variant calls using a standalone instance provided by George Wright^22^. The somatic variant calling and annotation pipelines are available as part of the LCR Modules suite of pipelines.

Structural variants were identified with an ensemble of GRIDSS2^52^ and Manta^48^ intersected with the BioConductor package StructuralVariantAnotation^53^. Copy number variants were identified with Battenberg^54^ for WGS or Sequenza^55^ for WES.

To identify immunoglobulin rearrangements, RNAseq data were analyzed with MiXCR^56,57^ with full contig assembly. IMGT Blast^58^ was used to annotate assembled contigs with IG genes.

### Quality Control

Genome coverage was estimated three ways: by calculating the theoretical coverage as read length multiplied by total unique reads divided by reference genome size (“Raw Coverage Estimate”), by calculating the average depth at all variant positions (“Mean Variant Depth”), or using Picard CollectWgsMetrics (“Mean Corrected Coverage”), which does not double-count the overlapping portions of read pairs^59^. Coverage of exome and targeted sequencing data were estimated with Picard CollectHsMetrics (Extended Data Figure 2).

### Clonal analysis

Shared variants were quantified in tumor pairs (using the first two DLBCL morphology time points for each patient) by matching variants on genome position and variant allele. The percent exclusive variants was calculated as the proportion of all variants in a tumor not shared with its paired biopsy. For clonal analysis, variant calls were subset to synonymous and non-synonymous variants in coding space and in regions commonly affected by aSHM^6^. A bed file of aSHM regions is available as part of the GAMBLR R package. Variants were clustered into subclones with PyClone-VI^60^ and phylogenetic relationships between subclones were reconstructed with its companion tool PhyClone. Battenberg results were provided to PyClone-VI for copy number and purity estimation.

To identify “trunk” variants frequently in the hypothetical common precursor cell (CPC) population, we leveraged patients with a branching evolution pattern in which all tumors had at least 25% exclusive mutations. *MYD88* L265P, *CREBBP* lysine acetyltransferase (KAT) domain missense mutations, *CD79B* Y197, *EZH2* Y646, and *NOTCH1/2* PEST domain truncating mutations were tallied separately from other mutations in each of those genes. Constrained genes were also identified in this subset of patients by identifying genes with exclusive mutations in two or more tumors from the same patient.

### Statistics and plotting

Categorical comparisons were made using Fisher’s exact test and comparisons of continuous variables were made using Wilcoxon rank sum tests. Linear models were used when comparing two or more continuous variables. For outcomes, P-values were derived from pairwise log-rank tests or cox models adjusted for available clinical information including age and international prognostic index (IPI). All statistics were performed and plots generated in R version 4.1.

## Extended Data

**Extended Data Figure 1.**
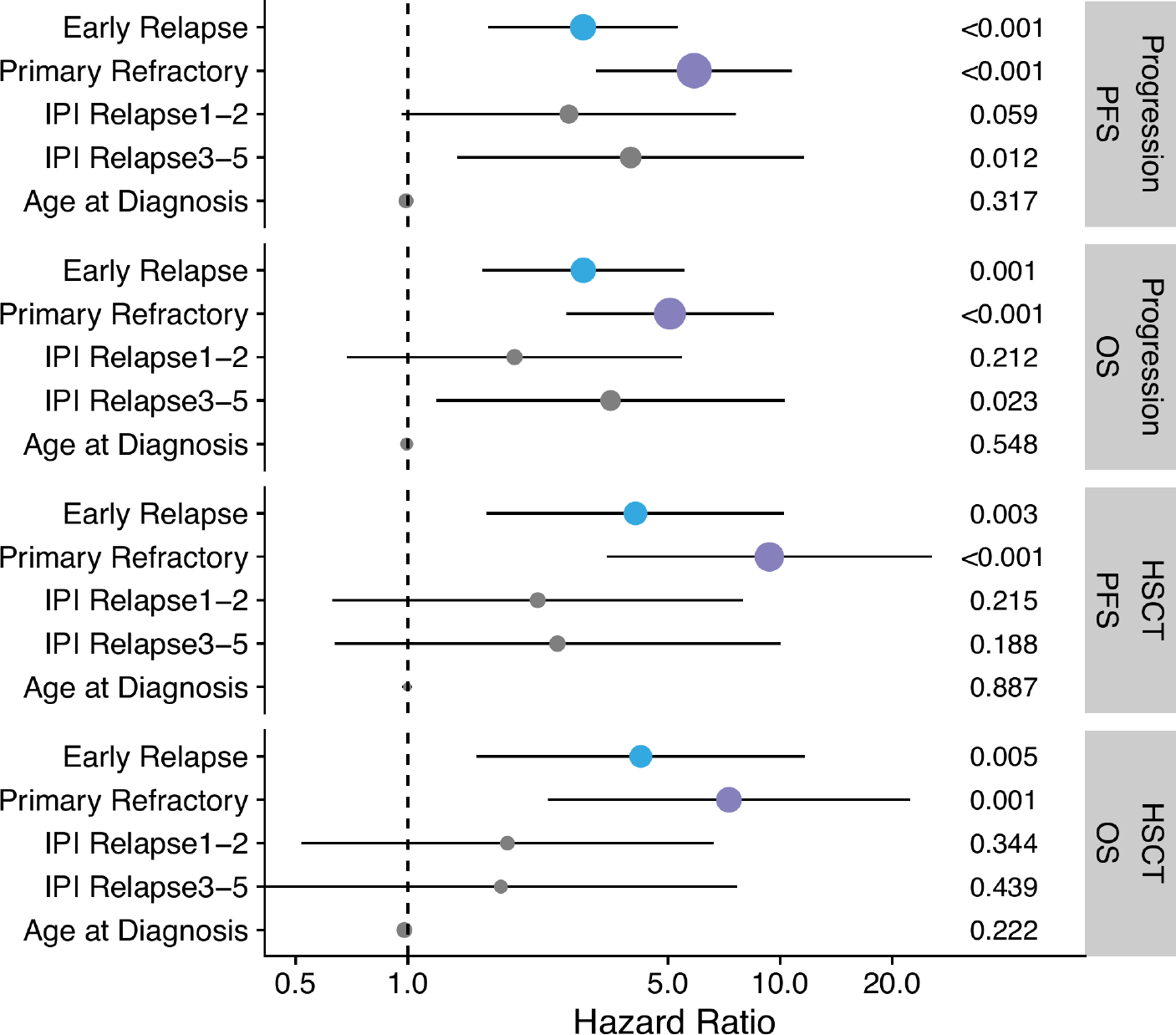
Hazard ratios and P-values from Cox models of OS and PFS from time of progression or time of transplant, adjusted for age at diagnosis, sex, and IPI at relapse.

**Extended Data Figure 2.**
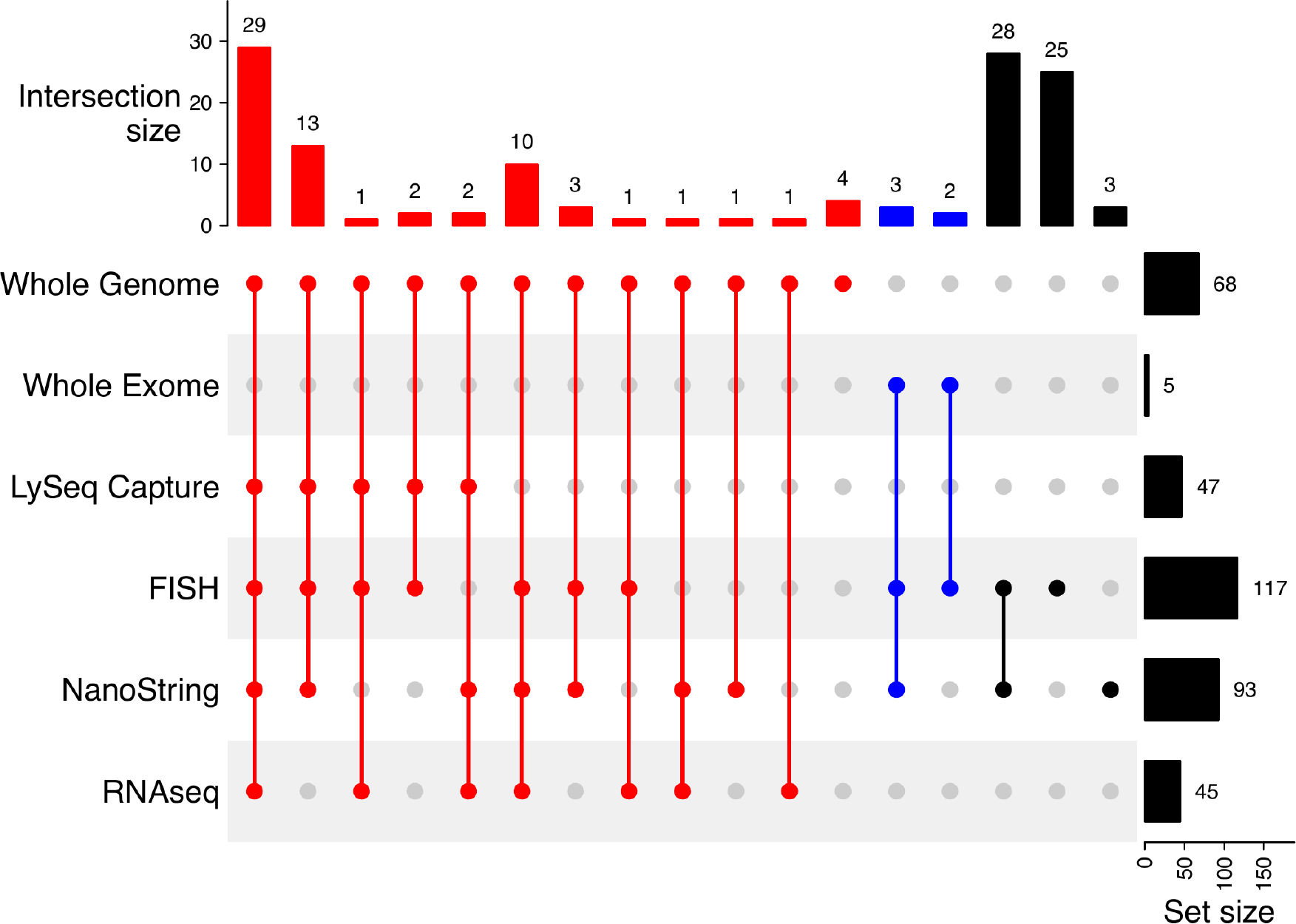
UpSet plot summarizing assays completed on multiple biopsies per patient.

**Extended Data Figure 3.**
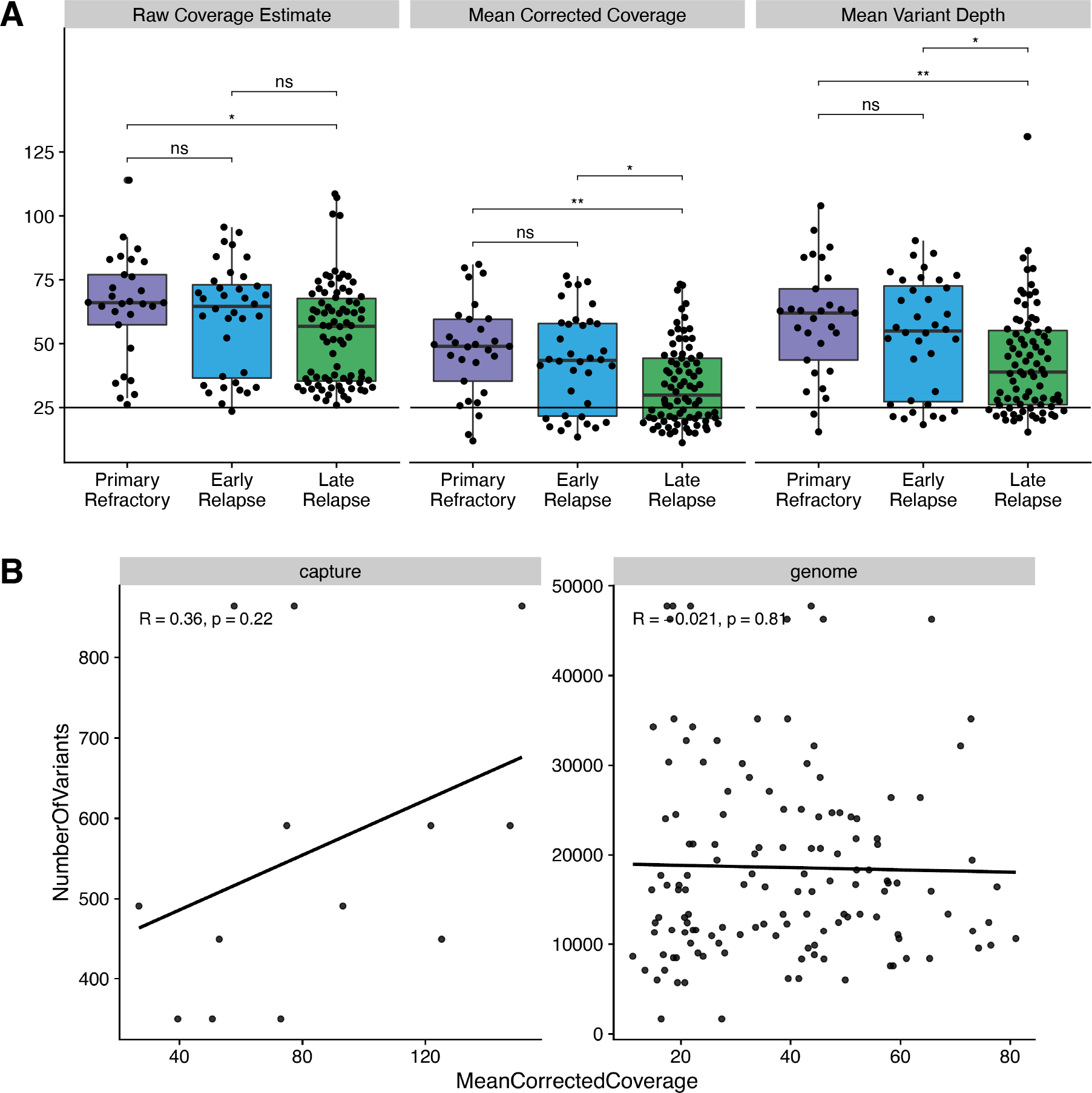
A. Raw coverage estimate, mean corrected coverage (not double-counting overlapping reads), and mean depth across each variant position across relapse timing categories. * P < 0.05; ** P < 0.01; ns not significant. B. Coverage vs. total mutation burden for exomes (“capture”) and genomes. R represents Pearson correlation coefficient.

**Extended Data Figure 4.**
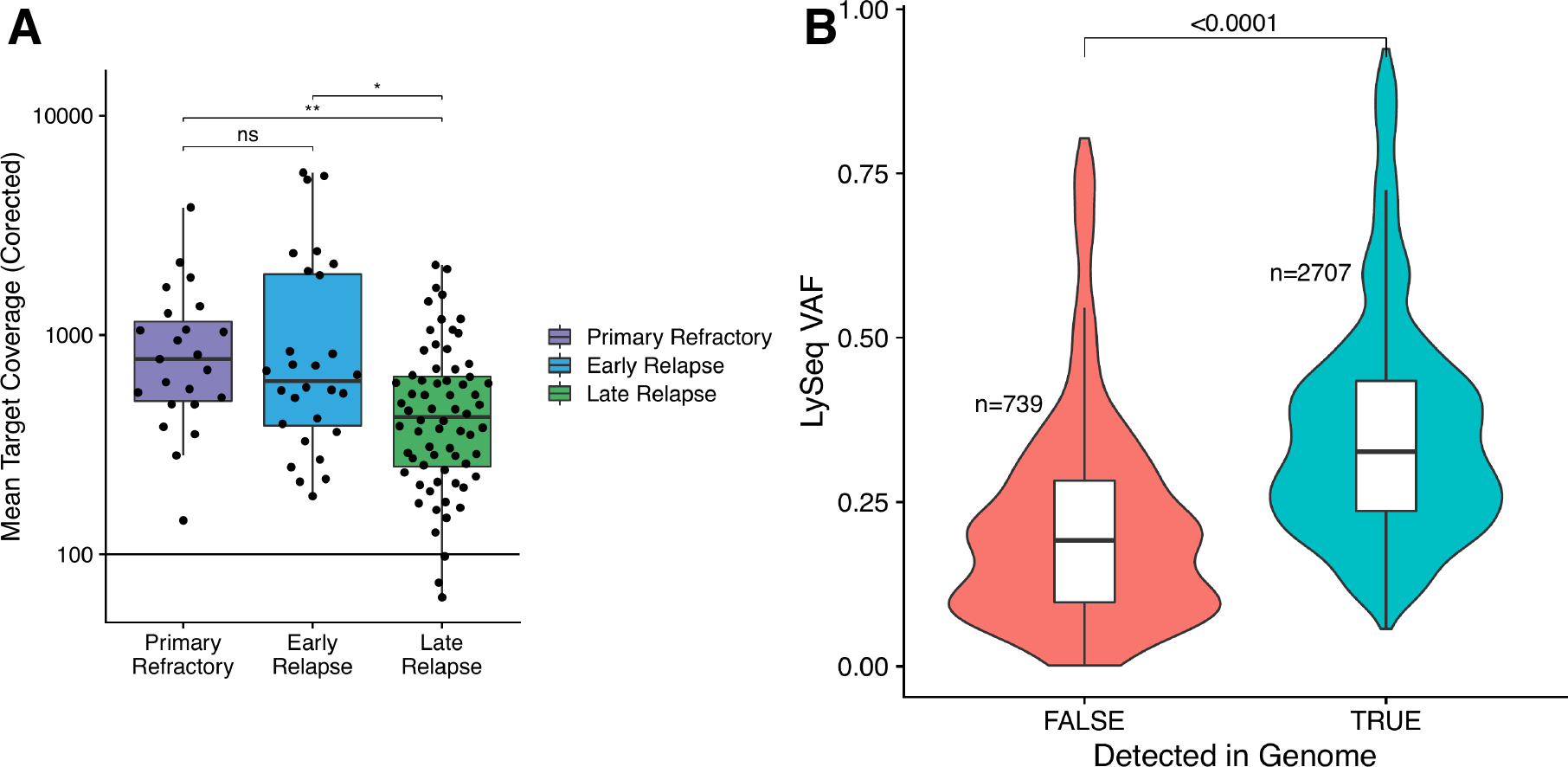
A. Mean corrected coverage across capture space for the LySeqST assay stratified by relapse timing category. * P < 0.05; ** P < 0.01; ns not significant. B. VAFs of variants identified with the LySeqST assay, stratified by whether or not a variant was called in the matching WGS data. Lower VAFs of variants not detected by WGS supports that LySeqST has enhanced sensitivity for sub-clonal variants that fall below the limit of detection of WGS.

**Extended Data Figure 5.**
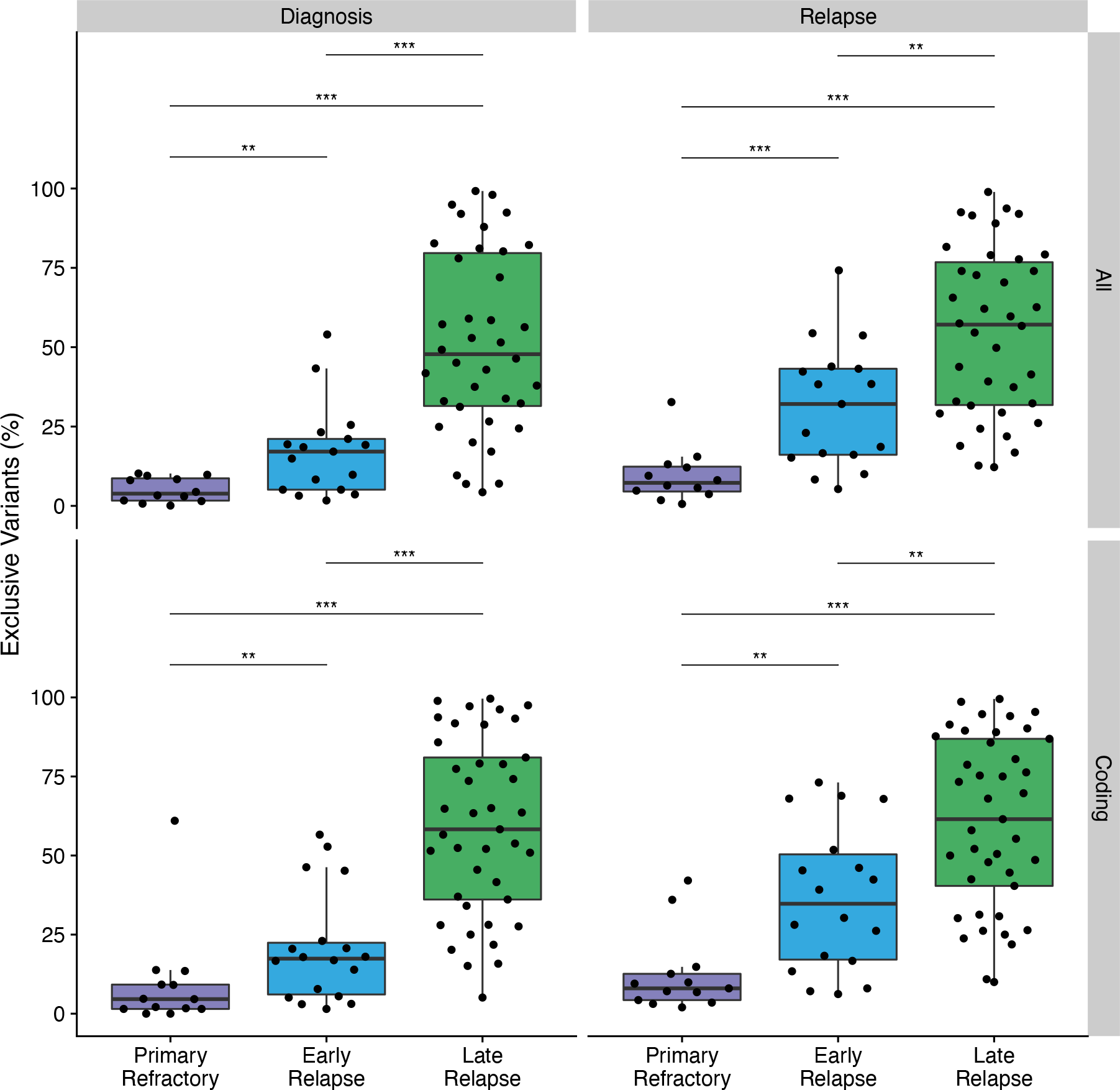
A. Percent exclusive variants per biopsy vs. relapse timing as a categorical variable. *** P < 0.001, ** P < 0.01, * P < 0.05. B. Correlation of percent exclusive variants with relapse timing, stratified by whether or not a patient ever had indolent disease. R represents Pearson corrrelation coefficient.

**Extended Data Figure 6.**
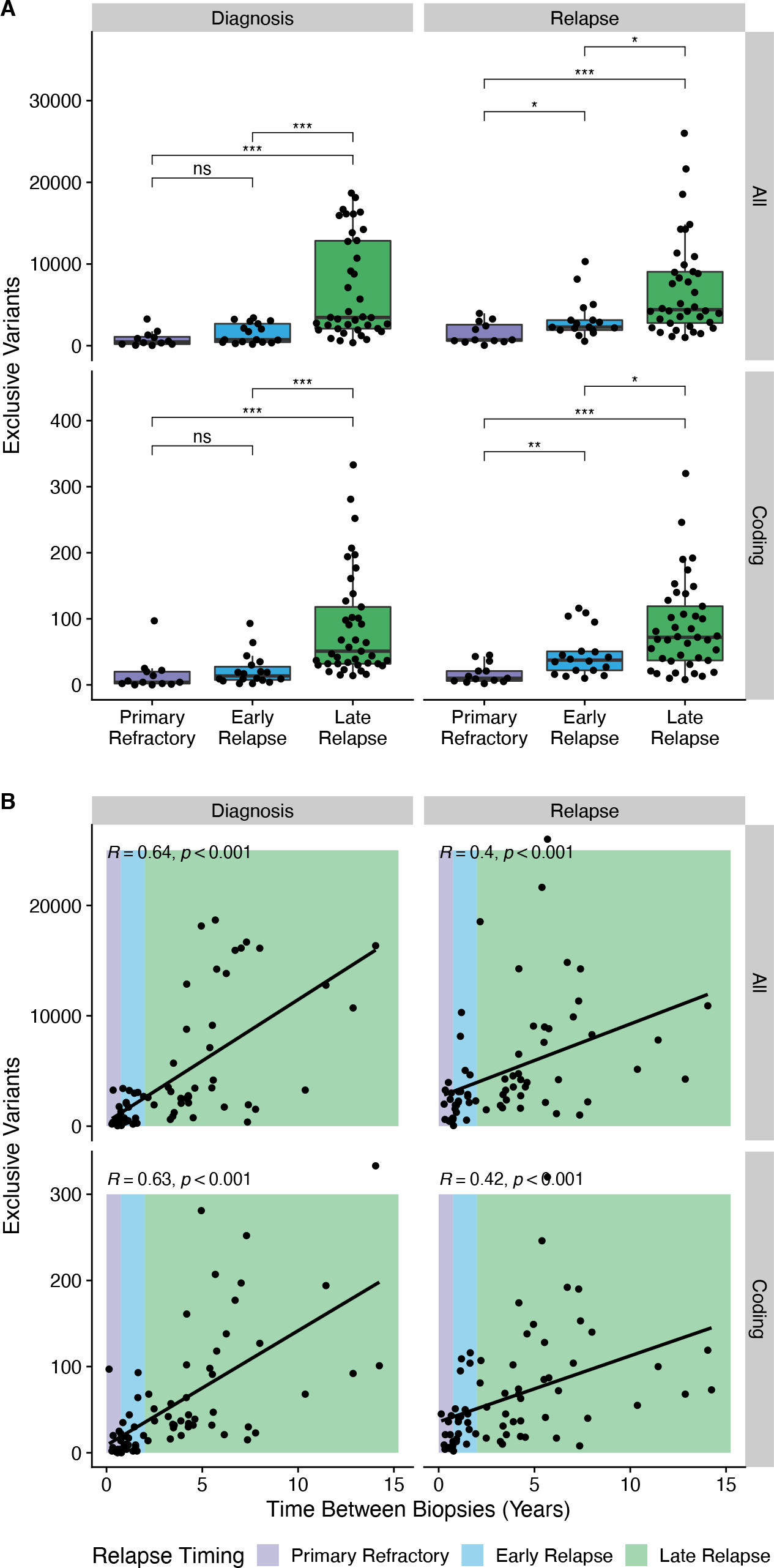
Relationship between absolute number of exclusive mutations per biopsy vs. relapse timing as a categorical (A) or continuous (B) variable.

**Extended Data Figure 7.**
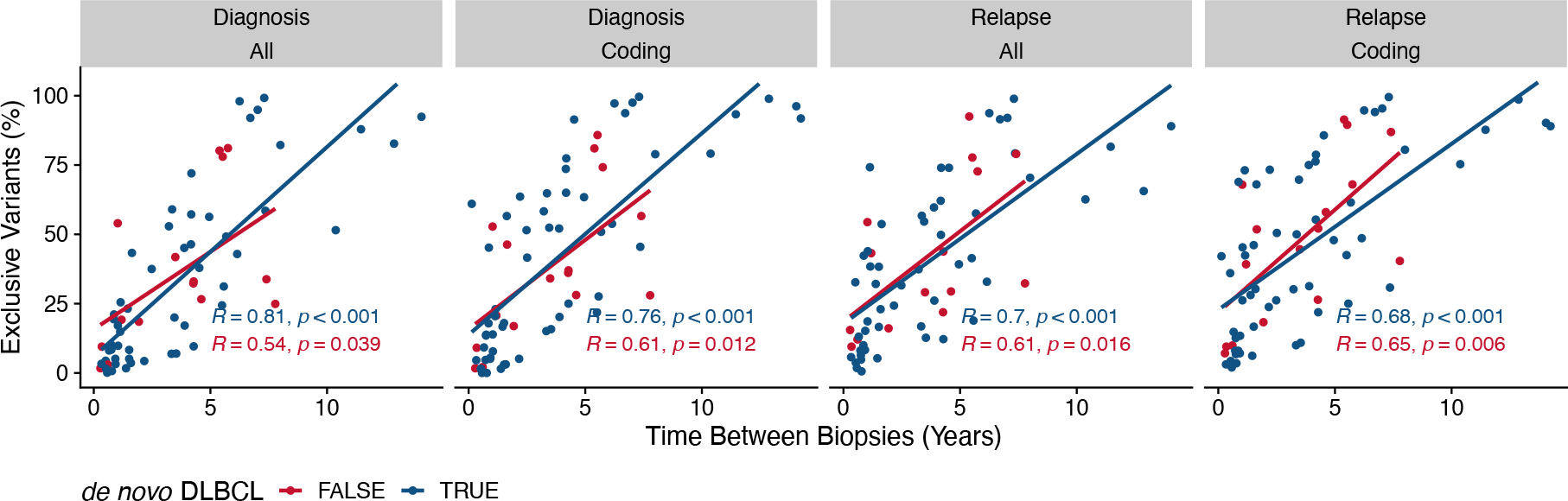
Correlation of percent exclusive variants with relapse timing, stratified by whether or not a patient had transformed disease. R represents Pearson corrrelation coefficient.

**Extended Data Figure 8.**
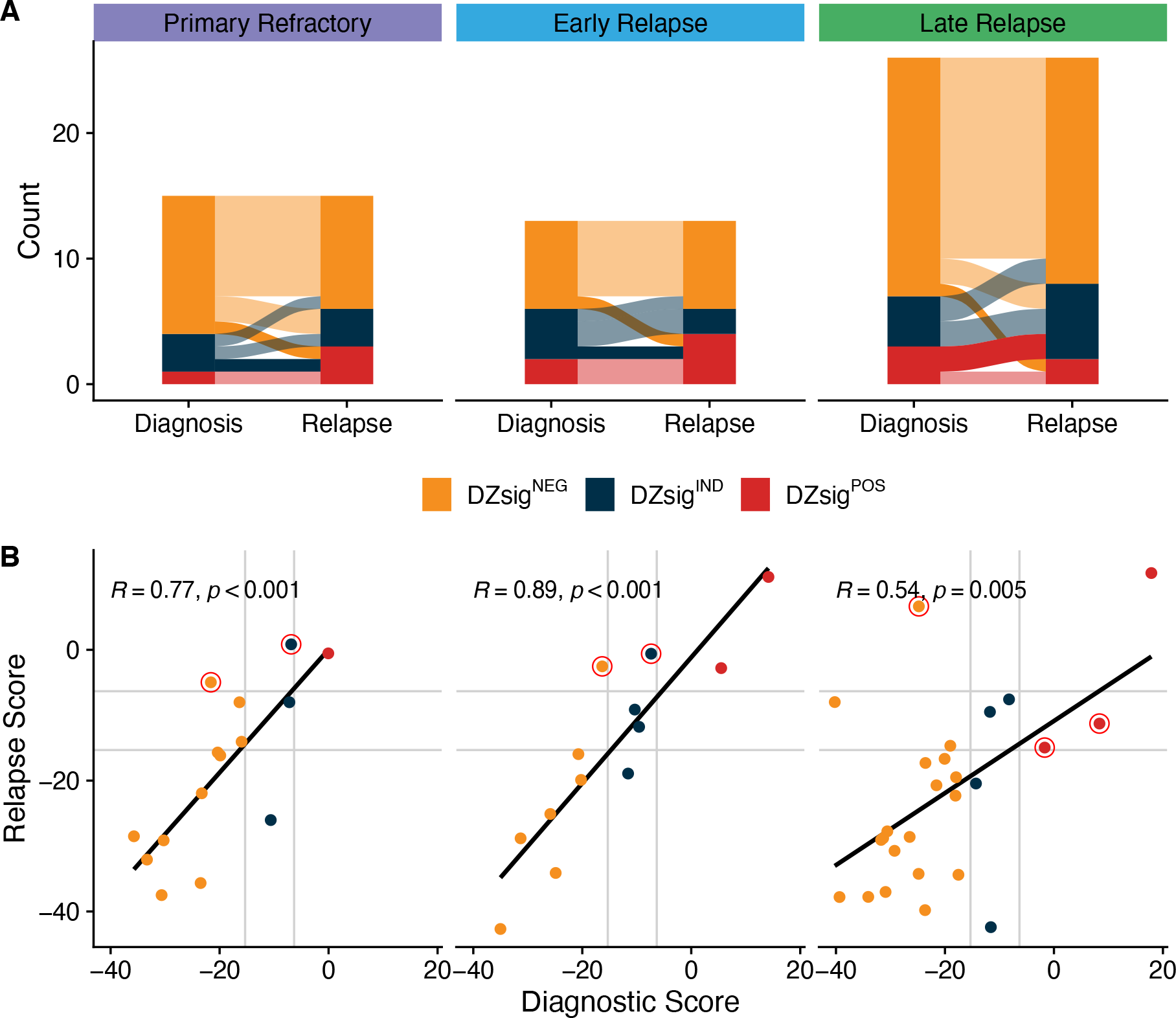
Comparison of DZsig classifications and scores. A. Alluvial plot highlighting switches from DZsigPOS to NEG or IND with opaque alluvia. B. Comparison of DLBCL90 DZsig scores between biopsies. R represents Pearson correlation. Only tumors classified as GCB or unclassified for COO were included in this analysis.

